# Laboratory Evaluation of *Cyclospora cayetanensis* in Wastewater: Expanding Pathogen Targets for Regulatory Public Health Surveillance

**DOI:** 10.64898/2026.10.08.26364996

**Authors:** Chi-Yu Lin, Dolores Sanchez Gonzalez, Rachel S. Poretsky

## Abstract

Wastewater-based epidemiology (WBE) has become a cornerstone of public health surveillance, yet routine regulatory monitoring remains largely focused on respiratory viruses and selecting antimicrobial resistant bacteria. Emerging foodborne parasites, such as *Cyclospora cayetanensis*, remain excluded from standard environmental surveillance panels despite increasing domestic outbreaks. This study presents a laboratory-based pilot protocol evaluating the feasibility of extracting and quantifying *C. cayetanensis* from urban wastewater across the Chicago metropolitan area. Using sample concentration via centrifugation and mechanical bead-beating, automated magnetic bead nucleic acid extraction, and digital PCR (dPCR), we demonstrate high-sensitivity detection in municipal influents and sewer nodes. These findings establish a proof-of-concept laboratory methodology to support the expansion of regulatory public health surveillance frameworks to include non-routine protozoan pathogens.

## 1. Introduction

*Cyclospora cayetanensis* is a waterborne and foodborne protozoan parasite responsible for cyclosporiasis— a gastrointestinal disease characterized by prolonged watery diarrhea, fatigue, and abdominal cramps^1^. Unlike viral pathogens, *C. cayetanensis* oocysts are environmental structures requiring days to weeks outside a host to sporulate and become infectious.

While public health agencies have successfully institutionalized wastewater monitoring for viral targets (e.g., SARS-CoV-2, Influenza, Poliovirus) and select multidrug-resistant bacteria, parasitic pathogens are rarely integrated into standard regulatory surveillance programs^2^. Clinical surveillance for *Cyclospora* is heavily constrained by underreporting, self-limiting mild infections, and specialized diagnostic requirements (such as modified acid-fast staining or multiplex PCR)^3^. Establishing robust laboratory workflows for detecting parasitic targets in wastewater is an essential first step toward demonstrating that protozoan pathogens can— and should—be incorporated into routine, regulated public health monitoring systems.

## 2. Current Situation in Our Society

In summer 2026, cyclosporiasis cases in the United States have increased steadily, with recurrent seasonal outbreaks in the Midwest linked to fresh produce supply chains^1^. Despite this growing burden, current municipal wastewater monitoring mandates do not encompass protozoan parasites.

The primary structural bottleneck in managing foodborne protozoan outbreaks is the delay in epidemiological detection. Clinical notification occurs weeks after exposure, by which time contaminated produce has typically cleared distribution networks. Furthermore, asymptomatic shedding in the community goes entirely uncaptured by clinical reporting. Incorporating *C. cayetanensis* into municipal surveillance frameworks would provide regulatory and health authorities with real-time, population-level baseline data, bridging the gap between clinical underreporting and proactive outbreak intervention.

## 3. Experimental Method

### 3.1 Environmental Sampling

Environmental samples were collected in August 2026 across the greater Chicago area to assess target recovery across diverse catchment scales. Sampling sites included community sewer nodes and primary raw influents at Metropolitan Water Reclamation District of Greater Chicago (MWRD) facilities. 24-hour composite and Moore swab samples were transported on ice to the laboratory for immediate processing.

### 3.2 Sample Concentration and DNA Extraction

To evaluate optimal recovery of resilient *C. cayetanensis* oocysts from complex matrices, an optimized laboratory workflow adapted from recent environmental pathogen monitoring frameworks was executed^4^:

1. **Centrifugation:** Wastewater samples were concentrated via centrifugation to pellet suspended solids and parasite oocysts.
2. **Mechanical Bead-Beating:** Pelleted material underwent mechanical disruption (bead-beating) to effectively rupture the resistant oocyst wall and liberate target genomic DNA.
3. **Automated Extraction:** Lysates were processed using MagMAX Viral/Pathogen Nucleic Acid Isolation reagents on the KingFisher Apex System (ThermoFisher) to maximize DNA yield while eliminating humic substances and PCR inhibitors.

### 3.3 Molecular Detection

Target DNA was detected and quantified using digital PCR (dPCR) via the QIAcuity Digital PCR System (QIAGEN). Digital PCR was selected for its superior resilience to matrix-derived inhibition and its ability to achieve absolute quantification of the target (copies/µL) without reliance on external calibration curves.

## 4. Preliminary Results

### 4.1 dPCR Detection of C. cayetanensis

Initial laboratory evaluation of 15 environmental wastewater samples across the Chicago metropolitan area confirmed robust target detection. *Cyclospora cayetanensis* was detected in **73.3%** (11/15) of environmental samples evaluated. Complete raw data for all tested samples are presented in Table 1, and their log-scale concentration distributions are compared in Figure 1.

**Table 1.** Raw Analytical Data for All Tested Samples.

| Table 1. Raw Analytical Data for All Tested Samples |  |  |  |  |
| --- | --- | --- | --- | --- |
| Sample ID | Target | Concentration (copies/μL) | Positive Partitions | Detection Status |
| A | Cyclospora | 1.218556 | 18 | Positive |
| B | Cyclospora | 0.406895 | 6 | Positive |
| C | Cyclospora | 0.504449 | 7 | Positive |
| D | Cyclospora | 1.278132 | 23 | Positive |
| E | Cyclospora | 0 | 0 | Negative |
| F | Cyclospora | 0.383894 | 5 | Positive |
| G | Cyclospora | 0.263376 | 5 | Positive |
| H | Cyclospora | 0 | 0 | Negative |
| I | Cyclospora | 0.353952 | 5 | Positive |
| J | Cyclospora | 0 | 0 | Negative |
| K | Cyclospora | 0.758416 | 15 | Positive |
| L | Cyclospora | 0.15401 | 3 | Positive |
| M | Cyclospora | 0.211362 | 4 | Positive |
| N | Cyclospora | 0 | 0 | Negative |
| O | Cyclospora | 4.706638 | 94 | Positive |

**Figure 1.**
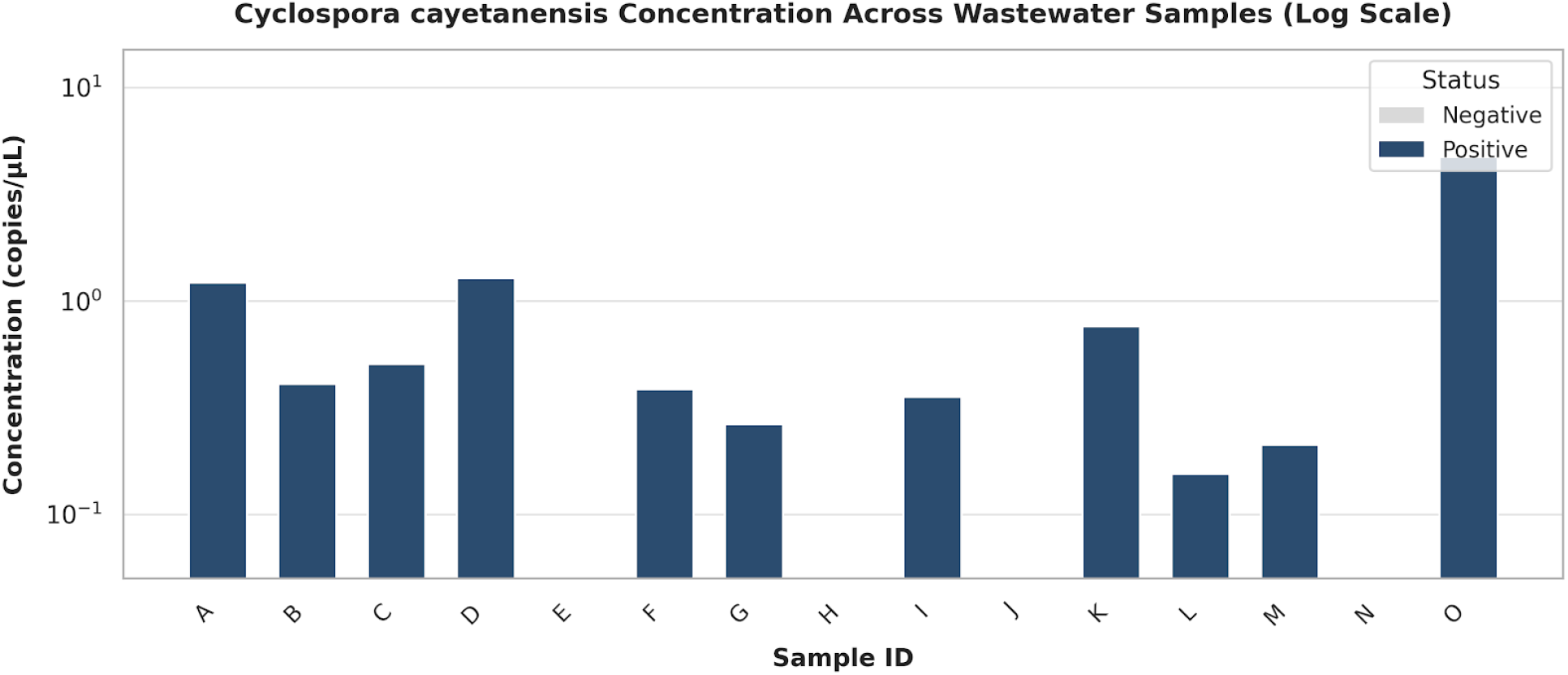
Cyclospora cayetanensis target concentration (copies/μL, logarithmic scale) across 15 environmental wastewater samples

Interpreting wastewater pathogen data requires accounting for multiple biological and environmental variables. Because shedding rates, magnitude, and duration exhibit significant inter-individual and inter-pathogen variability, translating wastewater concentrations into precise epidemiological metrics remains scientifically challenging^5^. Consequently, wastewater surveillance data should not be evaluated in isolation but should be integrated with traditional clinical and public health indicators to inform comprehensive community health assessments.

## 5. Discussion & Plan for Health Surveillance Advocating

Demonstrating reliable laboratory detection is the prerequisite for expanding routine public health monitoring. Moving forward, this work aims to advocate for formal regulatory integration through the following strategic initiatives:

- **Expanding Regulatory Target Panels:** Presenting methodological feasibility data to public health authorities (e.g., local/state health departments) to justify adding *C. cayetanensis* and other foodborne parasites to standardized wastewater monitoring mandates.
- **Establishing Threshold Baselines:** Conducting longitudinal benchmarking to establish background signal levels, enabling regulatory bodies to define actionable thresholds for public health warnings or epidemiological investigations.
- **Standardizing Laboratory Protocols:** Providing verified, high-throughput extraction (centrifugation + bead-beating) and dPCR protocols to reference labs to harmonize parasite surveillance across municipal regions.
- **Cross-Agency Collaboration:** Partnering with agricultural and food safety regulatory bodies to utilize wastewater parasite data as an early signal for regional food supply contamination events.
- **Genomic Sequencing:** Integrating genomic sequencing approaches, where feasible, to characterize detected targets, support source attribution, and improve understanding of pathogen diversity and transmission patterns.
- **Analyzing Temporal Dynamics:** Examining long-term detection trends over time to characterize temporal variation, seasonal fluctuations, and their potential public health implications to guide future research directions and targeted interventions.

## Declarations

## Ethical Approval

Not applicable. This study used wastewater samples and did not involve human subjects.

## Data Availability

All data generated or analyzed during this study are included in this article (see Table 1).

## References

1. Almeria, S., Cinar, H. N., & Dubey, J. P. (2019). Cyclospora cayetanensis and cyclosporiasis: An update. Microorganisms, 7(9), 317.

2. Bivins, A., North, D., Ahmad, A., et al. (2020). Wastewater-based epidemiology: Global collaborative to maximizecontributions in the fight against COVID-19. Environmental Science & Technology, 54(13), 7754–7757.

3. Centers for Disease Control and Prevention (CDC). (2023). Cyclosporiasis (Cyclospora Infection) - ealthcareProfessionals.

4. Poretsky, R., Sanchez Gonzalez, D., Horton, A., et al. (2025). Sewer monitoring for antimicrobial resistance genesand organisms at healthcare facilities. The Journal of Infectious Diseases, jiaf434. 10.1093/infdis/jiaf434

5. McClary-Gutierrez, J. S., Mattioli, M. C., Marcenac, P., et al. (2021). SARS-CoV-2 wastewater public healthreporting and communication: A review. Journal of Environmental Quality, 50(3), 614–628.

